# An Examination of Sustained Versus Transient Distressing Psychotic-Like Experiences Using Adolescent Brain Cognitive Development℠ Study Data

**DOI:** 10.1101/2020.11.10.20229229

**Authors:** Nicole R. Karcher, Rachel L. Loewy, Mark Savill, Shelli Avenevoli, Rebekah S. Huber, Carolina Makowski, Kenneth J. Sher, Deanna M. Barch

## Abstract

**Objective:** A potential distinguishing factor between more benign and transient psychotic-like experiences (PLEs) versus PLEs that predict risk for psychiatric disorders is whether the PLEs are sustained and distressing (sustained dPLEs). The current study examined associations of both sustained and transient dPLEs with relevant risk factors (e.g., cognition), use of mental health services, and functional correlates (e.g., school performance) in school-age children.

**Method:** The current study used three Adolescent Brain Cognitive Development℠ study data waves to create sustained dPLE (n=272), transient dPLE (n=244), and control (n=272) groups. Hierarchical linear models examined whether these groups differed in terms of use of mental health services, functional correlates, family history of mental disorders, other symptoms (e.g., parent-rated psychotic symptoms, internalizing, externalizing symptoms), environmental factors (e.g., adverse childhood events [ACEs]), cognitive functioning, developmental milestone delays, and neuroimaging indices.

**Results:** Several factors were more strongly associated with sustained versus transient dPLEs, including use of mental health services (for sustained vs. controls: d=0.38), drop in grades (d=--.30), other symptoms (i.e., parent-rated psychotic, bipolar, internalizing, externalizing, suicidality; 0.33>ds<0.88), ACEs (d=.36), and lower fluid and executive functioning cognitive scores (-0.31>ds<-0.41). For most risk factors, the sustained dPLEs group showed the greatest impairments, followed by the transient group, with the control group showing the least impairments.

**Conclusions:** These results have implications for understanding the pathogenesis of dPLEs, including indicating that several factors may distinguish transient from persisting dPLEs in children, including higher symptoms and ACEs, lower executive functioning scores, greater use of mental health services, and worsening school performance.

---

Psychotic-like experiences (PLEs), including nonclinical psychosis spectrum symptoms (e.g., perceptual abnormalities, mild delusional thoughts), are relatively common in school-age children, with ∼17% of 9-12-years-olds experiencing PLEs (1). Importantly, a subset of these children endorsing PLEs are at risk for conversion to psychotic disorders or other psychiatric disorders in adulthood (2). However, previous research has not yet elucidated what distinguishes those who later develop psychiatric disorders, including psychosis spectrum disorders, versus those who do not. This information would have critical implications for the future application of early identification and preventative interventions.

One potential distinguishing factor between more benign PLEs and those that transition to psychotic disorders is whether they are sustained (i.e., persisting over years) (3,4). Previous research indicates that sustained PLEs are generally associated with greater clinical relevance, including subsequent onset of psychotic disorders (3,5). Another potential distinguishing factor is the level of distress elicited by PLEs (6), as this may contribute to functional impairment. The relevance of distress is supported by studies showing that rather than frequency of psychosis spectrum symptoms, the associated distress predicts the onset of psychotic disorders (7). Furthermore, most studies examine cross-sectional estimates of PLEs, while only a limited number have examined PLE developmental trajectories (8), finding evidence for a sustained and a transient/relatively benign subgroup. However, no studies have specifically examined trajectories of distressing PLE*s*, as will be done in the current study.

PLEs in general are thought to arise as a result of a combination of genetic, environmental, and pathophysiological factors (e.g., disruptions in connectivity) (9,10). Several large cross-sectional datasets over the past two decades have examined the correlates of PLEs, finding associations with internalizing symptoms and externalizing symptoms (11), developmental impairments (12), and cognitive impairments (13), including reading (14), working memory (15), and processing speed (16) impairments. Of studies examining *trajectories* of PLEs, individuals with sustained PLEs tended to score higher on internalizing and externalizing symptoms (5), adverse childhood experiences (ACEs) (17,18), and developmental delays (18). Sustained PLEs are also associated with functional impairments and treatment seeking (3). However, a recent review did not find any consistent predictors emerging across studies (5). Furthermore, previous work has not examined sustained *distressing* PLEs (i.e., sustained dPLEs). In terms of associations with cross-sectional estimates of dPLEs, the Adolescent Brain Cognitive Developmentlil (ABCD) Study has found associations with a range of risk factors for psychopathology including psychosis, including family history of psychosis, other symptoms including internalizing, externalizing, bipolar, and suicidality symptoms, environmental risk factors including ACEs and exposure to deprivation, drug crime exposure, and lead exposure risk, cognitive impairments, developmental milestone delays, resting state functional connectivity (RSFC) impairments, and global structural MRI impairments (19-22). The current study examined whether, as predicted, sustained dPLEs are more strongly associated with deficits in the aforementioned risk factors, greater use of mental health services, and worse school and social functioning than transient dPLEs using data from 9-10-year-olds in the ABCD Study®.

## Methods

### Participants

The ABCD Study is a large-scale study tracking 9-10-years-olds recruited from 21 research sites across the United States. The current data release, ABCD Data Release 3.0 (DOI 10.15154/1519007) includes 3 waves of data: baseline (N=11,878), year 1 (N=11,235), and year 2 (N=6,571). These data were accessed from the National Institutes of Mental Health Data Archive (see Acknowledgments; see Supplement for study-wide exclusion details). As can be seen in Table 1, three groups were created using data from baseline, year 1, and year 2: 1) a *sustained dPLEs group* that had >=97.5 percentile PQ-BC distress scores for at least two waves of data, 2) a *transient dPLEs group* that had significantly elevated dPLEs (i.e., >=97.5 percentile PQ-BC distress score) in 1 wave and did *not* have significantly elevated scores (i.e., <=50 percentile PQ-BC) on the other waves (see Supplemental Table 1 for additional group membership details) (8), and 3) a *control group*, acting as a further comparison for the sustained and transient dPLEs groups, which had a stable low trajectory and included participants with no significantly elevated scores (i.e., <=50 percentile PQ-BC) on all ABCD waves (i.e., baseline, year 1, and year 2; note, group percentiles were re-normed at each wave). The control group was matched to the sustained dPLEs group on sex, age, and race/ethnicity. All measured risk factors (detailed below) were obtained at baseline.

**Table 1.** Group Characteristics^a^

| | Sustained dPLEs<br>(n=272) | Transient dPLEs<br>(n=244) | Controls (n=272) | 2-Tailed Test<br>Statistic (*t*, $\chi^2$) | *p*-<br>value |
| --- | --- | --- | --- | --- | --- |
| Sex (% female) | 48.2% | 56.6% | 48.2% | 4.75 | .09 |
| Race/Ethnicity |  |  |  |  |  |
| White | 37.5% | 38.9% | 37.5% | 0.15 | .93 |
| Black | 23.2% | 21.3% | 23.2% | 0.33 | .85 |
| Asian | 0.4% | 1.2% | 0.4% | 1.99 | .37 |
| Other | 11.0% | 11.5% | 11.0% | 0.03 | .98 |
| Hispanic | 27.9% | 27.0% | 27.9% | 0.07 | .97 |
| Age | 9.82 (0.61) | 9.92 (0.59) | 9.82 (0.61) | 0.31 | .76 |

Abbreviation: dPLE=distressing psychotic-like experiences.
<sup>a</sup>The control group was matched to the sustained dPLEs group in terms of propensity scores. HLMs were used to compare means across groups. $\chi^2$ Tests were used to compare ordinal/binary variables across groups.
<sup>b</sup>Means (SEs).

### Measures

All measures are described in detail within the Supplement.

#### Prodromal Questionnaire-Brief Child Version (PQ-BC)

Distress scores from the Prodromal Questionnaire-Brief Child Version (PQ-BC) (23), a 21-item self-report questionnaire, were used to create sustained, transient, and control groups.

#### Other Symptom and Functioning Measures

Modules from the Kiddie-Structured Assessment for Affective Disorders and Schizophrenia (K-SADS) for DSM-5 (24), including parent-rated psychotic symptoms (25), current bipolar symptoms, and externalizing symptoms (i.e current attention deficit hyperactivity disorder, oppositional defiant disorder, and conduct disorder symptom summations) (25), as well as child-rated internalizing symptoms (i.e., current depression and generalized anxiety disorder symptom summations), and suicidality (i.e., current suicidal ideation and behavior symptom summations) were used as measures of psychopathology.

Use of mental health services was measured by asking whether the youth has ever received mental health services. School performance was measured using KSADS questions regarding how well the youth does in school and whether there was a drop in grades over the past year. Social functioning was measured by asking the youth about number of friends.

History of psychotic disorder, depression, and mania in first-degree relatives was assessed using the Family History Assessment Module Screener (26), with each scored as either present or absent (family history of psychosis was an exclusionary criterion in the scoring of family history of depression and mania). Any history of psychotic disorders was scored as present if the participant had any first- or second-degree relatives with a history of psychotic disorder.

#### Neuropsychological Test Battery

The current study utilized uncorrected National Institutes of Health Toolbox Cognitive Battery scores from the 7 individual tests and fluid and crystallized composite scores, but all analyses include age and sex as covariates (27).

#### Developmental Milestones

The current study examined parent-reported motor (i.e., rolling over, sitting, and walking delays, subjective motor delays, clumsiness) and speech (i.e., delays in speaking first word, subjective speech delays) developmental milestone delays (28), including individual milestones and both motor and speech composite scores.

#### Environmental Risk Factors

Based on the child’s primary address coordinates, overall deprivation percentile scores, lead exposure risk, rate of poverty (defined as number of individuals living in poverty), drug crime exposure estimates were calculated. The current study also examined Adverse Childhood Experiences (ACEs), parent-rated perception of neighborhood safety, and number of years at current residence. The ACEs variable was defined as summations of parent-rated child experience of traumatic experiences from the K-SADS, a parent-rated question from the K-SADS about whether the child was bullied at school or in the neighborhood, and seven parent-rated questions of financial adversity from a demographic questionnaire (e.g., “Were [you] evicted from your home for not paying the rent or mortgage?”).

#### Structural MRI Measures

For the current study, structural MRI measures include total volume (intracranial, supratentorial, cortical, and subcortical) (29), surface area (30), and cortical thickness (31). All data were acquired on a 3T scanner (Siemens, General Electric, or Phillips) with a 32-channel head coil and completed T1-weighted and T2-weighted structural scans (1mm isotropic).

#### Resting State Functional Connectivity (RSFC)

Participants completed four 5-minute resting-state BOLD scans, with their eyes open and fixated on a crosshair. Consistent with previous research using the ABCD sample to examine associations with PLEs (22), we examined cingulo-opercular (CON) within-network connectivity, cingulo-parietal (CPAR) within-network connectivity, default mode (DMN) within-network connectivity, CON to cerebellar connectivity, and CPAR to cerebellar connectivity.

### Covariates

Every model included age, sex, and race/ethnicity (i.e., White, African American, Hispanic, Asian, Other) as covariates. However, aside from ACEs models, environmental risk factor models did not include race/ethnicity as covariates.^1^ Lastly, imaging analyses included scanner type as a covariate, with RSFC analyses additionally including average motion as a covariate.

### Statistical Analyses

The analyses used hierarchical linear models (HLMs) conducted using the R lme4 package (32) (multcomp package for multiple comparison analyses (33)). Variables that were zero-inflated (i.e., developmental milestones) were examined using zero-inflated generalized linear mixed models (glmmTMB package) (34). All analyses modeled family unit and research site as random intercepts. HLMs analyzed whether the groups (i.e., sustained, transient, controls) showed significant differences for each of the variables of interest. Pair-wise comparisons used to examine group differences were False Discovery Rate (FDR)-corrected across the three groups.

## Results

In general, the results detailed below follow a similar pattern, whereby across all factors examined, the sustained dPLEs group generally showed the greatest impairments, followed by the transient dPLEs group, with the control group showing the least evidence of impairment. Below we provide details about each domain.

### dPLEs

As can be seen in Table 2 and Figure 1, at every time point the sustained group showed greater dPLEs than both the transient and control groups. The transient group also showed greater dPLEs than the control group.

**Table 2.** Comparisons between Groups for Symptoms, Functional Correlates, Family History of Mental Illness, and Environmental Risk Factors^a^

|  | Sustained dPLEs (n=272) |  | Transient dPLEs (n=244) |  | Controls (n=272) |  | Sustained vs. Transient |  | Sustained vs. Controls |  | Transient vs. Control |  |
| --- | --- | --- | --- | --- | --- | --- | --- | --- | --- | --- | --- | --- |
|  | *M* | *SD* | *M* | *SD* | *M* | *SD* | *t* | *FDRp* | *t* | *FDRp* | *t* | *FDRp* |
| **dPLEs** |  |  |  |  |  |  |  |  |  |  |  |  |
| Baseline | 35.39 | 18.82 | 20.12 | 18.86 | 1.71 | 2.53 | **10.75** | **<.001** | **24.47** | **<.001** | **13.47** | **<.001** |
| Year 1 | 35.32 | 18.50 | 9.97 | 14.07 | 1.46 | 3.35 | **20.57** | **<.001** | **28.31** | **<.001** | **7.17** | **<.001** |
| Year 2 | 25.89 | 14.87 | 9.91 | 13.64 | 0.78 | 1.59 | **14.96** | **<.001** | **23.86** | **<.001** | **9.34** | **<.001** |
| **Use of Mental Health Services and Functional Correlates** |  |  |  |  |  |  |  |  |  |  |  |  |
| Use of Mental Health Services | 0.28 | 0.45 | 0.19 | 0.39 | 0.13 | 0.33 | **2.10** | **.04** | **4.36** | **<.001** | **2.20** | **.04** |
| School Performance | 3.30 | 0.74 | 3.39 | 0.69 | 3.62 | 0.54 | -1.35 | .18 | **-5.43** | **<.001** | **-3.96** | **<.001** |
| Drop in Grades | 0.18 | 0.39 | 0.10 | 0.30 | 0.08 | 0.27 | **2.99** | **.004** | **4.03** | **<.001** | 0.94 | .35 |
| Number of Friends | 23.66 | 25.01 | 21.92 | 20.57 | 20.61 | 18.39 | 1.00 | .48 | 1.54 | .37 | 0.51 | .61 |
| **Family History** |  |  |  |  |  |  |  |  |  |  |  |  |
| 1<sup>st</sup> Degree Psychosis | 0.04 | 0.20 | 0.04 | 0.20 | 0.02 | 0.15 | 0.43 | .66 | 1.28 | .61 | 0.83 | .61 |
| Any Psychosis | 0.17 | 0.38 | 0.10 | 0.30 | 0.09 | 0.29 | **2.65** | **.01** | **2.67** | **.01** | -0.04 | .97 |
| 1<sup>st</sup> Degree Depression | 0.35 | 0.48 | 0.29 | 0.45 | 0.24 | 0.43 | 1.04 | .30 | 2.32 | .06 | 1.32 | .28 |
| 1<sup>st</sup> Degree Mania | 0.07 | 0.25 | 0.08 | 0.27 | 0.03 | 0.18 | -0.09 | .93 | 0.19 | .93 | 0.27 | .93 |
| **Other Symptoms** |  |  |  |  |  |  |  |  |  |  |  |  |
| Parent-Rated Psychotic | 0.62 | 1.89 | 0.28 | 1.31 | 0.14 | 0.74 | **2.42** | **.02** | **3.82** | **<.001** | 1.30 | .20 |
| Parent-Rated Bipolar | 0.94 | 1.95 | 0.52 | 1.44 | 0.23 | 0.58 | **2.75** | **.009** | **5.42** | **<.001** | **2.55** | **.01** |
| Internalizing | 2.38 | 3.55 | 1.02 | 2.51 | 0.12 | 0.72 | **6.16** | **<.001** | **10.23** | **<.001** | **3.85** | **<.001** |
| Parent-Rated Externalizing | 6.57 | 9.73 | 3.68 | 7.54 | 1.80 | 4.75 | **3.64** | **<.001** | **6.84** | **<.001** | **3.07** | **.002** |
| Suicidality | 0.51 | 1.30 | 0.24 | 0.82 | 0.02 | 0.22 | **3.17** | **.002** | **5.73** | **<.001** | **2.45** | **.01** |
| **Environmental Risk Factors** |  |  |  |  |  |  |  |  |  |  |  |  |
| ACEs | 1.92 | 2.32 | 1.29 | 1.59 | 1.09 | 2.24 | **3.94** | **.001** | **4.74** | **<.001** | 1.15 | .25 |
| Overall Deprivation Percentile | 51.07 | 28.22 | 44.11 | 29.08 | 42.25 | 29.58 | 0.88 | .58 | 0.87 | .58 | 0.06 | .95 |
| Lead Exposure Risk | 5.72 | 3.21 | 5.73 | 3.07 | 5.34 | 3.09 | 0.10 | .92 | 0.44 | .92 | 0.38 | .92 |
| Rate of Poverty | 23.91 | 16.71 | 22.78 | 16.72 | 21.14 | 17.29 | 0.16 | .87 | 0.30 | .87 | 0.16 | .87 |
| Perceptions of Neighborhood Safety | 10.77 | 3.13 | 11.22 | 2.99 | 11.72 | 2.92 | -1.53 | .13 | **-3.52** | **.001** | -1.91 | .08 |
| Drug Crime Exposure | 6.77E+03 | 7.49E+03 | 6.43E+03 | 7.58E+03 | 5.74E+03 | 7.21E+03 | 0.30 | .77 | 1.59 | .32 | 1.25 | .32 |
| Years at Residence | 4.78 | 4.01 | 5.57 | 4.30 | 5.74 | 4.26 | -1.08 | .42 | -1.39 | .42 | -0.37 | .71 |

Abbreviations: $n$ =sample size; $M$ =mean; $SE$ =Standard Error; $t$ = $t$ -statistic; $FDRp$ =False Discovery Rate-corrected $p$ value; dPLEs=distressing psychotic-like experiences; ACEs=Adverse Childhood Experiences.
<sup>a</sup>Significant estimates in bold. Test statistics are 2-tailed.

**Figure 1.**
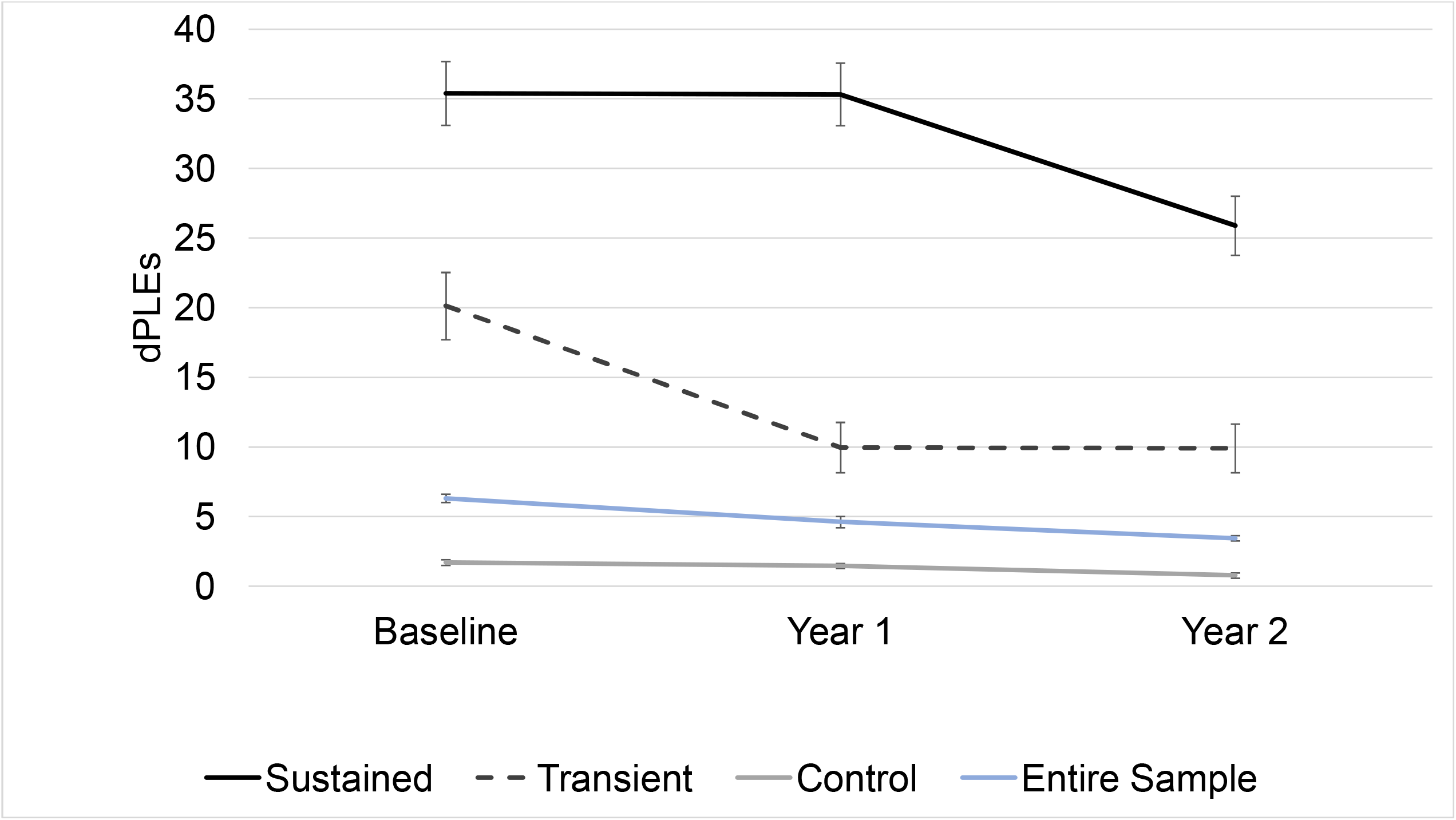
Average distressing psychotic-like experience (dPLE) scores for participants at each assessment wave (0=baseline; 1=year 1; 2=year 2) for each of the groups and for the entire sample. Error bars reflect 95% confidence intervals.

### Use of Mental Health Services and Functional Correlates

The sustained group showed greater use of mental health services and greater drop in grades compared to the transient dPLEs group (Table 2, Figure 2). In addition, both sustained and transient groups showed greater mental health services use and worse school performance compared to the control group, with the sustained dPLEs additionally showing a greater drop in grades compared to the control group.

**Figure 2.**
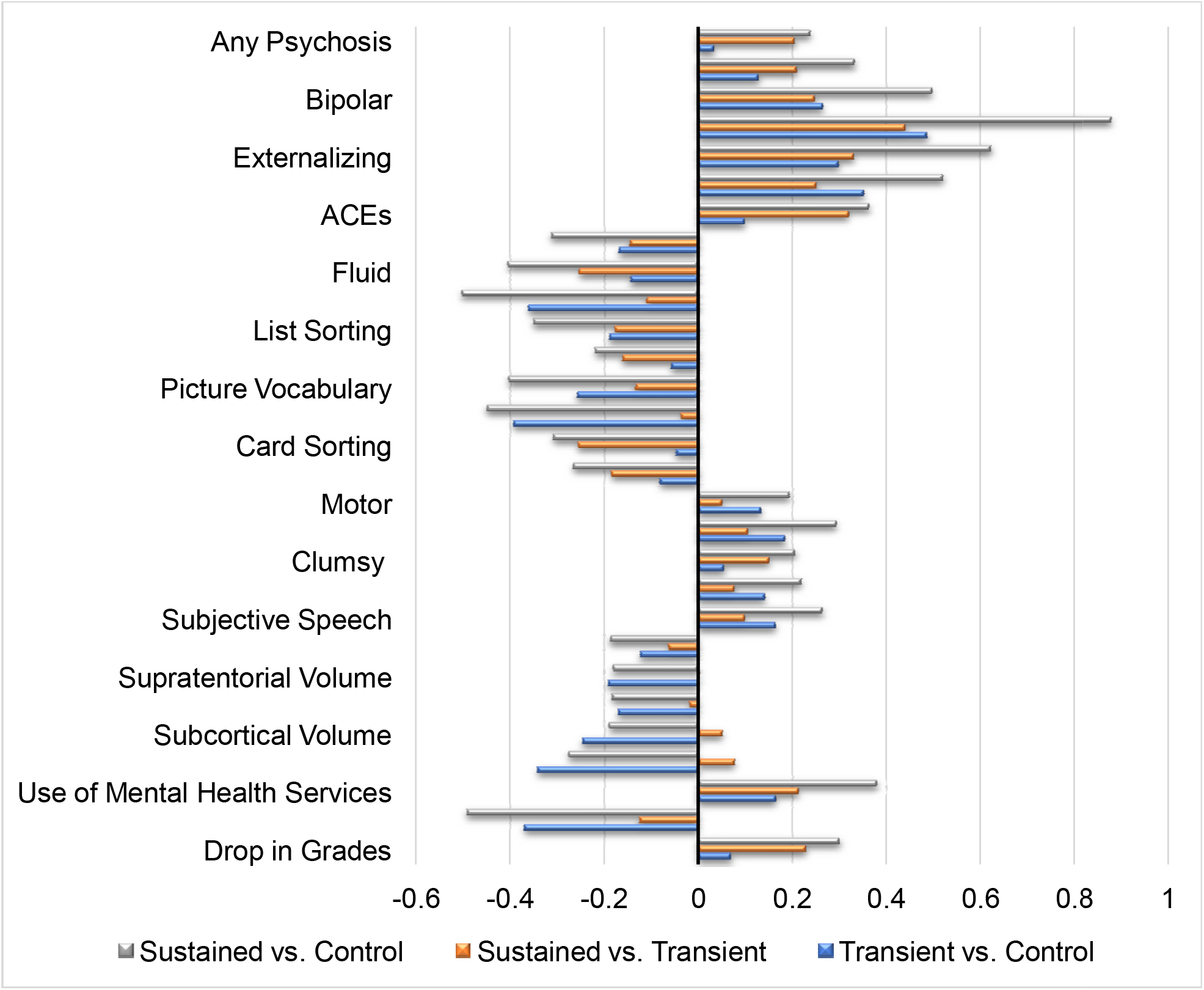
Effect sizes (Cohen’s D) for each of differences between the groups for risk factors with significant effects (Abbreviations: ACEs=Adverse Childhood Events; CON=cingulo-opercular).

### Family History of Psychosis

As can be seen in Table 2, although the sustained versus transient groups did not differ in number of first-degree relatives with a history of psychosis, the sustained dPLEs group showed a higher number of family members with a history of psychosis than either the transient or control group (Figure 2).

### Other Symptoms

At baseline, the sustained dPLEs group showed higher levels of all other symptoms compared to both the transient and control groups (i.e., parent-reported psychotic symptoms, bipolar, internalizing, externalizing, suicidality; see Table 2 and Figure 2). Notably, the transient dPLEs group did not differ from controls on parent-reported psychotic symptoms, although the transient group showed higher levels of bipolar, internalizing, externalizing, and suicidality symptoms than controls.

### Environmental Risk Factors

Even after including age, sex, and race/ethnicity as covariates, the sustained dPLEs group showed higher levels of ACEs than both the transient and control groups (Table 2, Figure 2). In terms of other environmental risk factors, the sustained dPLEs group showed lower parent-reported perception of neighborhood safety than the control group.

### Neuropsychological Test Performance

As can be seen in Table 3, the fluid composite showed a graded pattern, with the sustained dPLEs group showing lower scores than the transient dPLEs group, who in turn showed lower scores than controls. For the crystallized composite, both the sustained and transient dPLEs groups performed worse than controls but did not differ significantly from each other. In terms of individual tests, the sustained dPLEs group showed lower scores compared to both the transient dPLEs and control groups on the card sorting executive function task, with the transient and control groups not differing significantly. For the pattern comparison processing speed and picture sequence memory tasks, only the sustained dPLEs group showed lower scores compared to the control group (see Figure 2; note, the transient group showed a pattern of lower scores than controls, but not statistically significant). List sorting working memory, picture vocabulary, and reading test scores were significantly lower in both the sustained and transient dPLEs groups compared to controls but did not differ significantly across dPLEs groups.

**Table 3.** Comparisons between Groups for Neuropsychological Test Performance, Developmental Milestone Delays, and Neuroimaging Risk Factors^a^

|  | Sustained dPLEs<br>(n=272) |  | Transient dPLEs<br>(n=244) |  | Controls (n=272) |  | Sustained vs.<br>Transient |  | Sustained vs.<br>Controls |  | Transient vs.<br>Control |  |
| --- | --- | --- | --- | --- | --- | --- | --- | --- | --- | --- | --- | --- |
|  | *M* | SD | *M* | SD | *M* | SD | *t* | FDR<sub>p</sub> | *t* | FDR<sub>p</sub> | *t* | FDR<sub>p</sub> |
| Neuropsychological Test Performance |  |  |  |  |  |  |  |  |  |  |  |  |
| Fluid | 86.71 | 10.35 | 89.37 | 10.61 | 90.87 | 10.12 | **-2.50** | **.01** | **-5.19** | **<.001** | **-2.63** | **.01** |
| Crystallized | 83.12 | 6.12 | 83.85 | 7.02 | 86.27 | 6.38 | -0.46 | .65 | **-5.71** | **<.001** | **-5.22** | **<.001** |
| List Sorting | 91.89 | 12.41 | 93.98 | 11.05 | 96.15 | 11.91 | -1.73 | .08 | **-4.37** | **<.001** | **-2.58** | **.02** |
| Pattern Comparison | 84.49 | 14.93 | 86.86 | 14.36 | 87.69 | 14.11 | -1.52 | .19 | **-2.88** | **.01** | -1.32 | .19 |
| Picture Vocabulary | 80.80 | 6.71 | 81.77 | 7.72 | 83.80 | 8.11 | -0.94 | .35 | **-5.05** | **<.001** | **-4.09** | **<.001** |
| Reading | 88.45 | 6.78 | 88.72 | 7.24 | 91.35 | 6.10 | 0.21 | .83 | **-4.81** | **<.001** | **-4.96** | **<.001** |
| Flanker | 92.56 | 9.34 | 92.66 | 10.09 | 93.78 | 8.68 | 0.25 | .80 | -1.49 | .21 | -1.72 | .21 |
| Card Sorting | 88.71 | 10.97 | 91.42 | 10.19 | 91.89 | 9.62 | **-2.52** | **.02** | **-3.79** | **.001** | -1.20 | .23 |
| Picture | 99.21 | 11.93 | 101.45 | 12.25 | 102.47 | 12.57 | -1.60 | .16 | **-3.07** | **.007** | -1.40 | .16 |
| Developmental Milestone Delays |  |  |  |  |  |  |  |  |  |  |  |  |
| Motor | 0.51 | 0.84 | 0.47 | 0.89 | 0.36 | 0.71 | 0.95 | .34 | 1.94 | .16 | 0.95 | .34 |
| Speech | 0.36 | 0.63 | 0.30 | 0.60 | 0.19 | 0.48 | 0.93 | .35 | **3.18** | **.005** | **2.22** | **.04** |
| Clumsiness | 0.21 | 0.41 | 0.15 | 0.36 | 0.13 | 0.34 | 1.69 | .14 | **2.52** | **.04** | 0.78 | .43 |
| Roll Over | 0.07 | 0.26 | 0.09 | 0.29 | 0.07 | 0.25 | -1.02 | .46 | 0.35 | .73 | 1.38 | .46 |
| Sit Unassisted | 0.05 | 0.22 | 0.07 | 0.26 | 0.04 | 0.20 | -1.44 | .23 | 0.23 | .82 | 1.68 | .23 |
| Walk Unassisted | 0.05 | 0.21 | 0.05 | 0.22 | 0.03 | 0.18 | 0.09 | .93 | 0.89 | .64 | 0.79 | .64 |
| Subjective Motor<br>Delays | 0.13 | 0.34 | 0.10 | 0.30 | 0.09 | 0.28 | 0.84 | .57 | 1.42 | .47 | 0.57 | .57 |
| Speak First Word | 0.14 | 0.34 | 0.11 | 0.31 | 0.07 | 0.26 | 0.82 | .41 | **2.58** | **.03** | 1.73 | .2 |
| Subjective Speech<br>Delays | 0.22 | 0.42 | 0.18 | 0.39 | 0.13 | 0.33 | 0.83 | .41 | **2.63** | **.03** | 1.77 | .12 |
| Global Structural MRI |  |  |  |  |  |  |  |  |  |  |  |  |
| Surface Area | 1.83E+05 | 1.95E+04 | 1.82E+05 | 1.80E+04 | 1.85E+05 | 2.11E+04 | -0.16 | .88 | -1.94 | .12 | -1.77 | .12 |
| Cortical Thickness | 2.74E+00 | 1.18E-01 | 2.75E+00 | 1.16E-01 | 2.76E+00 | 1.13E-01 | -1.37 | .26 | **-2.41** | **.049** | -1.03 | .30 |
| Intracranial Volume | 1.46E+06 | 1.48E+05 | 1.47E+06 | 1.46E+05 | 1.50E+06 | 1.45E+05 | -0.28 | .78 | -2.34 | .06 | -2.05 | .06 |
| Supratentorial<br>Volume | 9.22E+05 | 9.22E+04 | 9.22E+05 | 9.22E+04 | 9.22E+05 | 9.22E+04 | 0.03 | .98 | **-2.54** | **.02** | **-2.56** | **.02** |
| Total Cortical<br>Volume | 5.76E+05 | 5.84E+04 | 5.77E+05 | 5.45E+04 | 5.87E+05 | 6.03E+04 | -0.45 | .65 | **-2.61** | **.03** | **-2.14** | **.0496** |
| Total Subcortical | 5.87E+04 | 5.12E+03 | 5.84E+04 | 4.54E+03 | 5.97E+04 | 5.74E+03 | 0.99 | .32 | **-2.94** | **.005** | **-3.93** | **<.001** |

| Volume |  |  |  |  |  |  |  |  |  |  |  |  |
| --- | --- | --- | --- | --- | --- | --- | --- | --- | --- | --- | --- | --- |
| Resting State Functional Connectivity |  |  |  |  |  |  |  |  |  |  |  |  |
| CON | 0.26 | 0.10 | 0.26 | 0.10 | 0.29 | 0.10 | 1.26 | .21 | **-2.14** | **.0496** | **-3.37** | **.002** |
| CPAR | 0.71 | 0.29 | 0.74 | 0.31 | 0.76 | 0.29 | -1.16 | .55 | -0.91 | .55 | 0.28 | .78 |
| DMN | 0.22 | 0.09 | 0.23 | 0.09 | 0.24 | 0.08 | -1.42 | .23 | -1.81 | .21 | -0.34 | .73 |
| CON-cerebellum | 0.05 | 0.09 | 0.05 | 0.11 | 0.04 | 0.07 | -1.06 | .29 | 1.17 | .29 | 2.22 | .08 |
| CPAR-cerebellum | -0.03 | 0.15 | -0.01 | 0.15 | 0.01 | 0.16 | -0.18 | 0.86 | -1.65 | .22 | -1.44 | .22 |

Abbreviations: n=sample size; *M*=mean; SE=Standard Error; t=t-statistic; FDRp=False Discovery Rate-corrected *p* value; CON=cingulo-opercular; CPAR=cingulo-parietal; DMN=default mode.
<sup>a</sup>Significant estimates in bold. Test statistics are 2-tailed.

### Developmental Milestone Delays

As can be seen in Table 3, the sustained dPLEs group showed greater speech developmental delays, greater delays in speaking their first word, and greater subjective speech delays than controls (see Figure 2), with the transient group also showing greater total speech developmental delays than controls. Additionally, the sustained dPLEs group showed greater parent-reported clumsiness compared to the control group.

### Global Structural Metrics

The sustained dPLEs group showed lower cortical thickness compared to the control group (see Table 3, Figure 2). In addition, both sustained and transient groups showed lower supratentorial, cortical, and subcortical volume compared to the control group, but did not differ significantly from each other.

### Resting State Functional Connectivity

The sustained and transient dPLEs group showed lower CON RSFC compared to controls (see Table 3; Figure 2).

## Discussion

The current study examined whether a sustained dPLEs group showing elevated dPLEs at multiple timepoints exhibited meaningful differences in terms of the degree of impairments across an array of domains compared to a transient dPLEs group. The current study found that several factors were more strongly associated with sustained versus transient dPLEs, including any family history of psychosis, greater parent-rated symptoms of psychosis, self-reported symptoms of suicidality, internalizing, and externalizing psychopathology, ACEs, use of mental health services, drop in grades, and both fluid composite and executive functioning test scores. For almost all risk factors, there was a noteworthy gradient in impairment, such that the sustained group showed the greatest impairments, followed by the transient group, with the control group showing the least impairments, consistent with some previous research (5,18).

Importantly, several findings emerged indicating that the sustained dPLEs group may exhibit more severe psychopathology and impairments in school performance compared to the other groups. The transient group showed greater mental health service use and worse school performance than controls, but the sustained group showed even greater severity of impairments in these domains, and additionally showed a greater drop in grades compared to both transient and control groups (for differences between the sustained and control groups, |Cohen’s ds| ranged from .38-.46, reflecting small-medium effect sizes). These data are consistent with the idea that clinicians may consider using persistence of dPLEs as a marker of identifying individuals most in need of evaluation and intervention.

The sustained dPLEs group showed a number of differences compared to both the transient and control groups in terms of family history of psychosis and other symptoms. First, the sustained group had a greater first- and second-degree family history of psychosis than the transient and control groups. The sustained dPLEs group also showed higher levels of both parent- and self-reported symptoms compared to both the transient dPLEs and control groups (although notably the transient dPLEs group also showed higher internalizing, externalizing, and suicidality symptoms compared to the control group), in line with previous work (5). Previous work has theorized that in psychosis-prone individuals, the presence of critical thoughts may influence PLEs to become increasingly negatively valenced. This may trigger increased internalizing and externalizing symptoms as well as increased rumination regarding these dPLEs, leading to persistence of these experiences (5). Our findings are potentially consistent with this framework. The higher parent-rated psychotic symptoms in the sustained dPLEs group compared to either the transient or control groups is critical caregiver-rated validation that this group is exhibiting more severe psychopathology, including significantly worse psychosis spectrum symptoms, and potentially pointing to greater caregiver symptom awareness with greater severity and longevity of PLEs.

In terms of environmental risk factors associated with the development of psychosis, sustained dPLEs were associated with higher ACEs compared to both the transient and control groups. This replicates previous work finding that sustained PLEs were associated with increased rates of trauma, including victimization (17,18). This finding is in line with theories that exposure to additional environmental risks, such as ACEs, can interact with genetic vulnerabilities to contribute to subclinical psychosis becoming initially distressing and sustained, potentially followed by a transition to a clinical psychotic state (9,10). The sustained dPLEs group also experienced lower perception of neighborhood safety compared to the control group. In terms of speculative mechanisms, previous work has theorized that increased chronic stressors cumulatively result in downstream neurobiological effects (e.g., dopamine sensitization, HPA axis dysfunction), potentially resulting in sustained dPLEs (4).

A similar graded pattern was found for neurocognition and developmental delays whereby the sustained dPLEs group generally showed the greatest impairments and the control group showed the least impairments. Several cognitive domains, including both fluid and crystallized intelligence composites, tests of working memory (listing sorting), picture vocabulary, reading, as well as speech development delays, showed differences for both sustained and transient dPLEs groups compared to controls. These impairments may be associated with dPLEs and/or psychopathology in general, as opposed to being specifically associated with more severe dPLEs (e.g., sustained dPLEs). For executive function (card sorting), processing speed (pattern comparison), episodic memory (picture sequence), time to speak first word, subjective speech delays, and clumsiness, the sustained dPLEs group showed greater impairments compared to the control group whereas the transient dPLEs group did not show a difference compared to the control group. The sustained dPLEs group showed lower fluid cognition and executive functioning scores compared to the transient group, perhaps indicating that these may be particularly important cognitive markers of persistence of PLEs. Overall, these findings are consistent with previous work finding language impairments (12,35), processing speed (16), motor abnormalities (36), and executive functioning (37) as potential important markers in the development of psychosis. The findings indicate that sustained dPLEs are associated with impairment in a range of cognitive functions, with evidence that executive functioning scores in particular may differentiate the sustained dPLEs from transient dPLEs and controls. Further, these findings may indicate that these risk factors are either predictors and/or consequences of greater severity of symptoms, such that neurocognition and developmental delays may partially account for presenting symptoms and problems and/or may reflect underlying anomalies in pathophysiology.

Several key findings emerged from the neuroimaging analyses. First, consistent with evidence that lower cortical thickness is associated with greater severity of psychosis spectrum symptoms, including the development of a psychosis spectrum disorder (38), the sustained dPLEs group showed lower cortical thickness compared to controls. Both the sustained and transient dPLEs groups showed lower cortical and subcortical volume than controls, consistent with previous research (39). Further, in terms of RSFC, both the sustained and transient dPLEs groups showed lower CON RSFC compared to the control group. Although speculative, alterations in cortical thickness may index greater impairment associated with sustained dPLEs, whereas alterations in subcortical volume and CON RSFC may be more associated with the psychosis spectrum phenotype more broadly.

The current study had several limitations and points to consider. First, the groups were created based on *a priori* definitions of groups membership. Based on the current study’s group definitions, it is possible that individuals that met transient criteria due to significant elevations only at year 2 (and not baseline and year 1) will meet criteria for the sustained dPLEs group in later waves of data collection. Further, it is possible that individuals that met sustained criteria at baseline and year 1 but not year 2 represent a ‘recovering’ group. However, the inclusion of these individuals likely only attenuated our results. Further, only including individuals that met criteria for the transient group at baseline and the sustained group at year 1 and year 2 generally produced similar results. Additionally, given that we were specifically interested in sustained versus transient dPLEs, we chose not to use a data-driven approach to create trajectories. Future research should continue to examine whether associations between sustained versus transient dPLEs remain consistent over time as sample sizes increase and more participants become eligible for these groups over time. Lastly, the measures used contain a mix of parent- and child-reports. For some measures (e.g., ACEs), parent report is limited by the parent’s awareness of the events and willingness to report (40).

The current research furthers our understanding of sustained versus transient dPLEs, finding evidence in support of the notion that dPLEs that persist over time are indeed associated with greater family history of psychosis, impairments in symptoms, use of mental health services, school functioning, and lower fluid and executive functioning scores. In addition, the study provided further evidence for ACEs as an important environmental risk factor in sustained dPLEs (3,4,9,10). There are several reasons why some risk factors may differentiate sustained from transient dPLEs and others do not. For example, risk factors differentiating sustained from transient dPLEs groups included factors that may be associated with the degree of distress and impairment currently experienced by the sustained dPLEs group (e.g., symptoms, use of mental health services, drop in grades). Furthermore, several risk factors, including developmental milestone delays, cognitive functioning, and several neuroimaging metrics showed gradient effects, with the sustained dPLE group showing larger effects than the transient group. These risk factors may further differentiate sustained versus transient dPLEs over time as the former group becomes increasingly condensed to those at risk for psychiatric disorders. Further, it is possible that impairments in several risk factors (e.g., cognition, neural impairments) may intensify over development, such as in adolescence. As future ABCD data releases become available, research will be able to examine risk factor trajectories of sustained dPLEs, including clinical and functional outcomes as these youth enter adolescence and young adulthood.

## Data Availability

Data used in the preparation of this article were obtained from the Adolescent Brain Cognitive Development (ABCD) Study (https://abcdstudy.org), held in the NIMH Data Archive (NDA).

## Acknowledgments

Data used in the preparation of this article were obtained from the Adolescent Brain Cognitive Development (ABCD) Study (https://abcdstudy.org), held in the NIMH Data Archive (NDA). This is a multisite, longitudinal study designed to recruit more than 10,000 children age 9-10 and follow them over 10 years into early adulthood. The ABCD Study is supported by the National Institutes of Health and additional federal partners under award numbers U01DA041022, U01DA041028, U01DA041048, U01DA041089, U01DA041106, U01DA041117, U01DA041120, U01DA041134, U01DA041148, U01DA041156, U01DA041174, U24DA041123, U24DA041147, U01DA041093, and U01DA041025. A full list of supporters is available at https://abcdstudy.org/federal-partners.html. A listing of participating sites and a complete listing of the study investigators can be found at https://abcdstudy.org/Consortium_Members.pdf. ABCD consortium investigators designed and implemented the study and/or provided data but did not necessarily participate in analysis or writing of this report. This manuscript reflects the views of the authors and may not reflect the opinions or views of the NIH or ABCD consortium investigators.

The ABCD data repository grows and changes over time. The ABCD data used in this report came from DOI 10.15154/1519007.

## Additional Contributions

We thank the families participating in the Adolescent Brain and Cognitive Development study.

## Footnotes

1 Race/ethnicity was not included as a covariate in these analyses, due to the all too frequent confounding of minority status with other relevant factors involved in the current study (e.g., deprivation, increased exposure to offenses, reduced access to resources)

