## Supplemental Methods for "An Examination of Sustained Versus Transient Distressing Psychotic-Like Experiences Using Adolescent Brain Cognitive Development℠ Study Data"

**Participants**

The ABCD study aimed to recruit a sample reflecting the demographic variation of the U.S. population, recruiting children using probability sampling from both public and private elementary schools. Study-wide exclusionary criteria were as follows: child not fluent in English, MRI contraindication (e.g., irremovable ferromagnetic implants or dental appliances, claustrophobia, pregnant), major neurological disorder, gestational age less than 28 weeks or birthweight less than 1,200 grams, history of traumatic brain injury, or had a current diagnosis of schizophrenia, autism spectrum disorder (moderate, severe), mental retardation/intellectual disability, or alcohol/substance use disorder (1,2). Parents provided written informed consent and all children provided assent.

**Prodromal Questionnaire-Brief Child Version (PQ-BC)**

Participants completed the Prodromal Questionnaire-Brief Child Version (PQ-BC) (3), modified for use with 9-10-year-olds based on a series of interviews assessing children’s understanding of the items, with a visual response scale included as a distress scale (4). Consistent with previous research (3,5), distress scores were calculated as the total number of endorsed questions weighted by level of distress [i.e., 0=no, 1=yes (but no distress), 2-6=yes (1+score on distress scale); range: 0-126].

**Other Symptom and Functioning Measures**

Modules from the validated and computerized parent-reported and child-reported Kiddie-Structured Assessment for Affective Disorders and Schizophrenia (KSADS) for DSM-5 (6-8) were used in current analyses as measures of psychopathology (1). The computerized self-administered parent and child versions of the KSADS show good to excellent concordance with the clinician-administered computerized KSADS (9). We examined a summation of parent-reported KSADS psychotic symptoms (6,7). We also examined internalizing symptoms (i.e., summations of current depression and generalized anxiety disorder symptoms) and current bipolar symptoms. We examined parent-reported externalizing symptoms using a KSADS composite of current attention deficit hyperactivity disorder, oppositional defiant disorder, and conduct disorder symptom summations (6,7). Suicidality was examined using summations of endorsements of current child-rated suicidal ideation (i.e., thinking of a method for a suicide attempt, suicidal thinking with intent to act, thinking of a specific suicidal plan) and suicidal behavior (i.e., self-injury with intent to die, self-injury and thinking you could die from behavior, made preparation for a suicide attempt, aborted or interrupted suicide attempts, the method of an actual suicide attempt, or a suicide attempt in which they thought they could die) from the KSADS.

Use of mental health services was measured at year 1 as whether the youth has ever received mental health services (1=yes; 0=no). School performance were measured at year 1 using K-SADS questions regarding school performance (indexed by asking the parent how well the youth does in school; rated from 4=very well to 1=failing) and whether there was a drop in grades over the past year (1=yes, 0=no). Social functioning was measured by asking how many friends the youth has. This measure was winsorized (to +3 SD) to minimize the influence of extreme values.

**Neuropsychological Test Battery**

Participants completed all tests within the National Institutes of Health Toolbox Cognitive Battery (NIHTB-CB) (10-12). The NIHTB-CB consists of 7 tasks, grouped into two composite scores. The fluid composite consists of Flanker Inhibitory Control and Attention, List Sorting Working Memory, Dimensional Change Card Sort, Pattern Comparison Processing Speed and Picture Sequence Memory. The crystallized composite consists of Picture Vocabulary and Oral Reading Recognition test (see (11) for descriptions of individual NIHTB-CB tests). The current study utilized uncorrected NIHTB-CB scores, but all analyses include age and sex as covariates.

**Developmental Milestones**

The parent assessment battery included questions of motor and speech developmental milestone delays (13-16). The motor delays composite was coded as the summation of delays in attaining motor milestones [i.e., rolling over (delayed=6 months or later), sitting (delayed=after 9 months), walking (delayed=after 18 months), parent-reported concern regarding motor development (parents were asked to compare their child’s development to that of other children: 0=earlier, 1=average, 2=later), and parent-reported current child clumsiness (16) and scored from 0=not true, 1=somewhat or sometimes true, and 2=very true or often true]. The speech delays composite was coded as the summation of a delay in speaking first word (delayed=after 12 months) and parent-reported concern regarding speech development (0=earlier, 1=average, 2=later). All motor and speech milestones were scored from 0=achieved within a typical timeframe or 1=delayed.

**Environmental Risk Factors**

Perception of neighborhood safety was calculated as a summation of three parent-rated questions (i.e., “I feel safe walking in my neighborhood, day or night”; “Violence is not a problem in my neighborhood”; “My neighborhood is safe from crime”; each was rated on a scale from 1-5, 1=strongly disagree, 5=strongly agree). A number of environmental risk factors were retrieved based on the child’s primary address coordinates. First, drug crime exposure information was obtained from the Uniform Crime Report from FBI, compiled by Inter-university Consortium for Political and Social Research (17), averaged from 2010-2012 to create stable county-level estimates of exposure to drug offenses. This measure was winsorized (to +3 SD) to minimize the influence of extreme values. Second, overall deprivation was defined as the Area Deprivation Index (ADI) national percentile scores, calculated from the 2011-2015 American Community Survey 5-year summary (18). Third, we examined the proportion of individuals living in poverty (-125% of poverty level) and number of years at current residence. Fourth, estimates of lead exposure risk were obtained by first geocoding the participant’s address at the census tract-level and then calculating risk scores based on data obtained from vox.com (https://www.vox.com/a/lead-exposure-risk-map). Estimated lead exposure risk scores (1-10, 10 being the most at risk) were calculated using proportion of individuals living in poverty and average age of the home.

**Structural MRI Measures**

Structural neuroimaging processing was completed using FreeSurfer version 5.3.0 through standardized processing pipelines (19). Participants that did not pass FreeSurfer Quality Control measure (i.e., at least one T1 scan that passed all quality control metrics) were excluded from analyses (n= 2). Cortical reconstruction and volumetric segmentation was performed by the ABCD Data Acquisition and Integration Core using the FreeSurfer image analysis suite (<http://surfer.nmr.mgh.harvard.edu/>). This pre-processing includes removal of non-brain tissue using a hybrid watershed/surface deformation procedure (20), automated Talairach transformation, segmentation of the subcortical white matter and deep gray matter volumetric structures, intensity normalization, tessellation of the gray/white matter boundary, automated topology correction, and surface deformation following intensity gradients (21). Images were registered to an atlas, which was based on individual cortical folding patterns to match cortical geometry across subjects.

**Resting State Functional Connectivity (RSFC)**

Resting state images were acquired in the axial plane using an EPI sequence. Other resting-state image parameters varied by 3T scanner and have been previously detailed (<https://abcdstudy.org/images/Protocol_Imaging_Sequences.pdf>) (22). The data analysis pipeline has also been detailed previously (23). Briefly, the cingulo-opercular within-network connectivity, cingulo-parietal within-network connectivity, default mode within-network connectivity, CON to cerebellar connectivity, and CPAR to cerebellar connectivity were derived from regions of interest (ROIs) within functionally-defined parcellations (i.e., Gordon networks; (24)) and subcortical ROIs (i.e., cerebellum; (25)). The Fisher Z-transform of the correlation values were examined. Participants were removed from analyses in the current study for not have at least one resting state scan that passed quality assurance criteria (n=10).

| Supplemental Table 1. Additional Group Membership Details | | | |
| --- | --- | --- | --- |
|  | Baseline | Year 1 | Year 2 |
| Sustained Group |  |  |  |
| N=49 |  |  |  |
| N=76 |  |  |  |
| N=43 |  |  |  |
| N=68 |  |  |  |
| N=36 |  |  |  |
| Transient Group |  |  |  |
| N=117 |  |  |  |
| N=57 |  |  |  |
| N=70 |  |  |  |
| = met group membership criteria based on data at this/these timepoint(s); = currently missing Year 2 assessment; note transient group membership was contingent on having data at baseline, year 1, and year 2. | | | |
